# Heterogeneous evolution of SARS-CoV-2 seroprevalence in school-age children: Results from the school-based cohort study Ciao Corona in November-December 2021 in the canton of Zurich

**DOI:** 10.1101/2022.05.31.22275814

**Authors:** Sarah R Haile, Alessia Raineri, Sonja Rueegg, Thomas Radtke, Agne Ulyte, Milo A Puhan, Susi Kriemler

**Affiliations:** Department of Epidemiology, Epidemiology, Biostatistics and Prevention Institute (EBPI), University of Zurich, Hirschengraben 84, 8001 Zurich

**Author notes:** Correspondence: Prof. Susi Kriemler, Epidemiology, Biostatistics and Prevention Institute (EBPI), University of Zurich, Hirschengraben 84, CH-8001 Zürich.

**Keywords:** SARS-CoV-2, seroprevalence, children, adolescents, school-based

## Abstract

**Background:** Much remains unknown regarding the evolution of SARS-CoV-2 seroprevalence and variability in seropositive children in districts, schools, and classes as only a few school-based cohort studies exist. Vaccination of children, initiated at different times for different age groups, adds additional complexity to understand how seroprevalence developed in the school aged population.

**Aim:** We investigated the evolution of SARS-CoV-2 seroprevalence in children and its variability in districts, schools, and classes in Switzerland from June/July 2020 to November/December 2021.

**Methods:** In this school-based cohort study, SARS-CoV-2 antibodies were measured in primary and secondary school children from randomly selected schools in the canton of Zurich in October/November 2020, March/April 2021, and November/December 2021. Seroprevalence was estimated using Bayesian logistic regression to adjust for test sensitivity and specificity. Variability of seroprevalence between school classes was expressed as maximum minus minimum sero-prevalence in a class and summarized as median (interquartile range).

**Results:** 1875 children from 287 classes in 43 schools were tested, with median age 12 (range 6-17), 51% 12+ vaccinated. Seroprevalence increased from 5.6% (95% CrI: 3.5-7.6%) to 31.1% (27.0-36.1%) in unvaccinated children, and 46.4% (42.6-50.9%) in all children (including vaccinated). Earlier in the pandemic, seropositivity rates in primary schools were similar to or slightly higher (<5%) than those in secondary schools, but by late 2021, primary schools had 12.3% (44.3%) lower seroprevalence for unvaccinated (all) subjects. Variability in seroprevalence among districts and schools increased more than twofold over time, and in classes from 11% (7-17%) to 40% (22-49%).

**Conclusion:** Seroprevalence in children increased greatly, especially in 2021 following introduction of vaccines. Variability in seroprevalence was high and increased substantially over time, suggesting complex transmission chains.

Trial Registration: ClinicalTrials.gov NCT04448717

## Introduction

More than two years into the SARS-CoV-2 global pandemic, it remains unclear to what extent transmission takes place in schools.^1^ The predominant opinion currently is that children of all ages appear to be equally susceptible to SARS-CoV-2 infection compared to adults and that transmission in children occurs primarily in the community children live, including at school, at home and in other settings. Fortunately, symptomatic or severe disease, hospitalization and death are much less common in children.^1-3^ This pattern holds true even under the higher transmissibility and dominance of the delta and omicron variants of concern (VOC) over other SARS-CoV-2 strains.^4-6^ Yet, studies have shown an elevated risk of SARS-CoV-2 infection for adults living in households with children attending schools in-person,^7 8^ especially for higher school grades,^8^ although this was not a consistent finding.^9^ Moreover, the number of reported outbreaks in school settings after the summer break of 2021 compared to earlier times has increased, in part due to the greater transmissibility of the delta and omicron VOC, but possibly also due to higher testing rates including repetitive pool testing in the school setting.^10 11^ The infection risk may also have varied depending on control measures in schools such as wearing masks, social distancing, hygiene, symptomatic or repetitive testing, and the vaccination status of families, peers and teachers.^8 12^ Overall, several studies documented a low probability of children getting infected within schools under the delta VOC,^13^ as reported also during the pre-delta period of the pandemic.^14-18^

The rate of previous natural infection in children can be estimated from seroprevalence studies and vary between countries and populations. Several studies show that seroprevalence has risen sharply up to 15-42% in children by summer 2021,^19-22^ and is expected to be even higher with the emergence of delta and omicron and the parallel increase in vaccination rates among children, adolescents, and adults. Most countries, including Switzerland, started vaccination in adolescents above 12 years from summer 2021 on, while vaccine was not approved for the 5-to 11-year-old group until late 2021 (in Switzerland mid-December 2021). The variability of vaccination rates among parents, teachers, and children below and above 12 years combined with the ever-changing transmissibility of new variants, differences in mitigation measures at school as well as in the community, and testing attitudes make surveillance of infection rates in school-age children a major challenge. Specifically, more evidence is needed regarding absolute levels of seroprevalence.

The Ciao Corona study examines SARS-CoV-2 seroprevalence among children in primary and secondary schools in one of the largest cantons of Switzerland.^23^ In this school-based prospective cohort study, four rounds of antibody testing have thus far been performed in June-July 2020 (T1),^24^ October-November 2020 (T2),^17^ March-April 2021 (T3)^16^ and November-December 2021 (T4) to assess the proportion of seropositive children and adolescents within cantonal districts, and by school, school level and class. While seroprevalence increased from 1.5 to 16.4% from T1 to T3, clustering of seropositive children within school classes was low and mostly reflected community transmission.^16^ Up to T3, seroprevalence in primary school was similar to or slightly higher (T2: primary 5.6% vs secondary 5.7%; T3: 19.5% vs 15.1%) than in secondary schools, possibly due to closer contact with their parents.^16 17^

The aim of this study was to describe the evolution of seroprevalence in children and adolescents from randomly selected schools of the canton of Zurich from October 2020 to December 2021, and to assess changes and variability of seropositive children within and across districts, schools, school levels and classes in our cohort. Due to low seroprevalences and recent school closure that may have led to infections more from households than from school in June-July 2020 and generally a lower positive predictive value in a low seroprevalence setting, we do not report T1 results here.

## Materials and Methods

The protocol for this school-based cohort study has previously been reported (ClinicalTrials.gov identifier: NCT04448717),^23^ as well as the results of the first three rounds of Ciao Corona testing.^16 17 24^ Ciao Corona, as part of the Swiss-wide research network Corona Immunitas,^25^ examines a randomly selected cohort of public and private schools and classes in the canton of Zurich, Switzerland. With 1.5 million inhabitants, the canton of Zurich is largest of 26 cantons in Switzerland by population and is home to a linguistically and ethnically diverse population in both urban and rural settings. Similar daily incidence of diagnosed SARS-CoV-2 cases until December 2021 in the canton of Zurich and Switzerland [Supplementary Material] document that the canton of Zurich is quite representative for Switzerland as a whole. The study was approved by the Ethics Committee of the Canton of Zurich, Switzerland (2020-01336). All participants provided written informed consent before being enrolled in the study.

After an initial lockdown period during which schools were closed from March to May 2020 across Switzerland, children have been physically in school without interruption. Preventive measures such as hygiene and social distancing rules, mask requirements were implemented in public and private schools according to cantonal rules, but with some variation within and among cantons. Children were required to stay home if they had a fever or other than minor cold symptoms. School personnel were required to wear masks starting in October 2020, while secondary school children (7th-9th grades) wore masks from November 2020, and the older primary school children (4th-6th grades) in early months of 2021, as well as again from December 2021. School-specific contact tracing was introduced in August 2020, where testing and quarantine recommendations depended on the situation. If at least two children were simultaneously tested positive in a class, then the whole class would be quarantined (existing policy from May 2020 to Feb 2022). However, if children were wearing masks, then quarantine was restricted to close contacts. Some schools started to participate in weekly pooled PCR testing in Spring 2021, with optional participation from children. By the time of T4 testing, approximately 80% of participating schools took part in repetitive testing, with variable participation rates of individual children, but at least 80% in most schools. In the case of a positive pool of up to 10 students, each child in that pool had a second individual test, and students remained at school but wore masks until the results were available. In the case of 3 or more positive cases in a class, negative tested children who participated in pool testing could continue to attend school, while children who did not participate in repetitive testing were then required to quarantine for 10 days.

### Population

As described elsewhere,^23^ public and private primary schools in the canton of Zurich were randomly selected in May 2020, and the geographically closest secondary school was also invited. The 55 participating schools (among them 2 private schools) were distributed in each of the 12 geographic districts proportional to the population size. Within participating schools, classes were randomly selected, stratified by school level: lower level (grades 1-3, age 6-9), middle level (grades 4-6, age 9-12), and upper level (grades 7-9, age 12-16). The aim was to invite at least 3 classes, with at least 40 children in each school level at a school. The invited sample is representative of the school-age population in the canton of Zurich.

Eligible children and adolescents (hereafter, children) in the selected classes could participate in any of the testing rounds and were reinvited to later testing periods. In the fourth round of testing, from November to December 2021, some schools declined to continue, reducing the total number of schools included from 55 to 43. Additional classes within the 43 participating schools were invited with the aim of obtaining a similar sample size to previous rounds. This resulted in 71 classes with only new children at T4 and 119 classes with a mix of new children at T4 or from previous rounds and 97 classes with only previously tested children. The main exclusion criterion was having a suspected or confirmed SARS-CoV-2 infection at the time of testing, which precluded attendance at school.

### Serological Testing

Venous blood samples were collected at schools Oct 26 – Nov 19, 2020 (T2), Mar 15 – Apr 16, 2021 (T3) and Nov 15 to Dec 14, 2021 (T4). Blood samples were analyzed using the Sensitive Anti-SARS-CoV-2 Spike Trimer Immunoglobulin Serological (SenASTrIS) test.^26^ The test uses Luminex technology to detect IgG and IgA antibodies binding to the entire trimeric S protein of SARS-CoV-2 and with demonstrated 94.0% sensitivity and 99.2% specificity for testing of IgG. It should be noted that previous publications of the Ciao Corona study reported results of a different Luminex-based test (e.g., ABCORA 2.3 binding assay), and therefore they will vary slightly from what is reported here.

At T4, two different definitions of seroprevalence were considered: a) Seroprevalence in tested children without documented vaccination, or b) Seroprevalence in all tested children (including those reporting vaccination). Because subjects who were both infected and vaccinated are not included in the first definition, seroprevalence will be underestimated. The true infection rate will be somewhere between the two definitions.

### Statistical Analysis

Statistical analysis included descriptive statistics and Bayesian hierarchical modelling to estimate seroprevalence. The Bayesian approach allowed to account for the sensitivity and specificity of the SARS-CoV-2 antibody test and the hierarchical structure of cohort (individual and school levels). The model (Bayesian logistic regression) was adjusted for participants’ grade and geographic district of the school and included random effects for school levels. We applied poststratification weights, which adjusted for the total population size of the specific school level and the geographic district. The model and weighting procedure are described in detail elsewhere.^24^

Variability in seroprevalence among districts, communities, schools, school level and classes was examined using variance partition coefficient (VPC) which describes the proportion of total variation that can be attributed to within unitvariability.^25^ With the introduction of the covid-19 vaccine to this age group in Switzerland, it is no longer possible to examine clustering as we have done in previous analyses of this study. Official statistics of SARS-CoV-2 infections in children aged 7-17 years of age in the canton of Zurich were retrieved to calculate the cumulative incidence of diagnosed SARS-CoV-2 cases by T2 to T4, and to compare with the proportion of seropositive children by Nov 6, 2020 (median time point of T2), March 29, 2021 (T3) and Nov 29, 2021 (T4).^27^

Socio-economic status of the school was measured with a composite measure *Social Index (SI)*, which reflects socioeconomic status (e.g., unemployment, immigrant population) of the location of the school and is provided by the Educational Directorate of Canton Zurich. Scores range from 100 to 120 with lower scores indicating less disadvantaged schools. All statistical analysis was performed using the R programming language,^28^ and Bayesian models were fit using the RSTAN package.^29^

## Results

Participant characteristics for T2, T3 and T4 are presented in Table 1. Among 930 participants aged 12 or older, 51% (n = 472) had been vaccinated at least 2 weeks prior to T4 serological testing. Among seropositive children, 50% in primary and 91% in secondary school were vaccinated. Of the 55 schools participating prior to T4, 43 continued to participate (Table 2). The total number of included classes increased from 275 to 287, and the median number of classes per school increased from 5 to 7 (range 2 to 10). The number of participants per school remained fairly constant, but participation at the class level was lower at T4 than during previous rounds. Socioeconomic status was comparable between schools that did participate and those that no longer participated in T4 (median 105 among schools not participating vs 107 among participating schools, p = 0.33).

**Table 1:** Characteristics of study participants over time.

|  | Oct-Nov 2020 | Mar-Apr 2021 | Nov-Dec 2021 |
| --- | --- | --- | --- |
|  | T2 | T3 | T4 |
| N total | 2500 | 2450 | 1875 |
| N (%) primary school<br>(grades 1-6) | 1594 (64%) | 1606 (66%) | 1050 (56%) |
| N (%) secondary school<br>(grades 7-9) | 906 (36%) | 837 (34%) | 825 (44%) |
| Age (median, range) | 12 (7 – 17) | 12 (7 – 17) | 12 (7 – 17) |
| Sex (n, % male) | 1211 (48%) | 1166 (48%) | 882 (47%) |
| Socio-economic status |  |  |  |
| High (n, %) | 1621 (70%) | 1616 (72%) | 1211 (73%) |
| Low-medium (n, %) | 687 (30%) | 633 (28%) | 443 (27%) |
| Vaccinated, aged 12+ | 0 | 0 | 472 / 930 (51%) |
| Seroprevalence adjusted* |  |  |  |
| infected only | 5.6% (3.5 to 7.6) | 18.4% (15.1 to 21.9) | 31.1% (27.0 to 36.1) |
| vaccinated ± infected |  |  | 46.4% (42.6 to 50.9) |
| Ratio of diagnosed to<br>seropositive | 1: 5.8 | 1: 3.5 | 1:3.5 |

Numbers denote participants (%). Seroprevalence denote means (95% credible interval). Socio-economic state was based on parental educational level with high denoting at least one parent with preparatory high school or university. \*Adjusted for school level, sex, and district, as well as test sensitivity and specificity. Seroprevalence due to infection only describes the total number of infected children with SARS-CoV-2 antibodies out of all unvaccinated children. Similarly, seroprevalence due to vaccination and/or infection is the total number of children with SARS-CoV-2 antibodies, regardless of vaccination status, out of all tested children.

**Table 2:** Participation rates and school-level characteristics of the Ciao Corona study at T2, T3 and T4.

|  | **Oct-Nov 2020** | **Mar-Apr 2021** | **Nov-Dec 2021** |
| --- | --- | --- | --- |
|  | **T2** | **T3** | **T4** |
| Number of schools | 55 | 55 | 43 |
| Number of classes | 276 | 275 | 287 |
| Median number of classes per school (min - max) [IQR] | 5 (2 – 10)<br>[3 – 6] | 5 (2 – 10)<br>[3 – 6] | 8 (3 - 18)<br>[5 – 8] |
| Median number of participants per school (min – max) [IQR] | 41 (13 – 101)<br>[31 – 56] | 37 (15 – 102)<br>[30 – 59] | 39 (16 - 94)<br>[27 – 58] |
| Median number of participants per class (min – max) [IQR] | 9 (1 - 20)<br>[5 – 12] | 9 (1 - 21)<br>[5 – 12] | 6 (1 - 17)<br>[2 - 8] |
| % participation within a class, median (min - max) [IQR] | 47% (5% - 94%)<br>[30% - 62%] | 50% (4% - 96%)<br>[32% - 63%] | 33% (4% - 89%)<br>[22% - 47%] |
| Number of classes with 5 or more participants | 222 | 221 | 203 |

### Evolution of seroprevalence

Seroprevalence at T2 was 5.6% (95% credible interval [CrI] 3.5 to 7.6%) and at T3 18.4% (15.1 to 21.9%) (Table 1). At T4, it was 31.1% (27.0 to 36.1) in non-vaccinated children and 46.4% (95% CrI 42.6 to 50.9) in all children (including vaccinated). Based on an overall PCR-positivity rate of 133 positive tests per 1000 inhabitants in the canton of Zurich aged 6-17 years, we estimate a ratio of diagnosed to seropositive children in late 2021 of 1: 3.5 which was similar to March – April 2021 (T3), though T2 had a higher proportion of children undiagnosed, 1: 5.8.

At T2 seroprevalence was similar across school levels (primary 5.5% (3.4 to 7.8%) vs secondary 5.6% (2.8 to 8.7%), while at T3, primary school children had a higher seroprevalence (19.5% (16.0 to 23.7)) than secondary school children (15.1% (10.7 to 19.6)). At T4 when counting only children with SARS-CoV-2 antibodies due to infection, and excluding vaccinated subjects, primary students had lower seroprevalence than secondary school students (28.7% vs 41.0%). However, when including both infected and vaccinated participants, secondary school students had an average seroprevalence of 75.8% (95% CrI 69.6 to 82.4), compared with 31.5% (27.1 to 36.1) among primary school students (grades 1-6) (Table 1).

### Variability of seroprevalence within and across districts, schools, and school classes

District level seroprevalence ranged from 30.4% to 88.5% at T4 (Figure 1a-c) documenting a substantial increase in variability over both T2 (2.1% to 18.0%) and T3 (11.1% to 27.2%). At T2 and T3, we observed only small differences between the school-level seroprevalence rates of primary and secondary schools (T2: primary 5.6% vs secondary 5.7%; T3: 19.5% vs 15.1%) (Figure 1a-c). However, by T4 considering the full sample (e.g. infected and/or vaccinated), all secondary schools had higher seroprevalence than all but one of the primary schools (Figure 1d-f). Within schools there was large variation between seroprevalence of classes (Figure 1g-i). At T2, median between-class variability was 11% (IQR 7% to 17%), at T3, 24% (17% to 37%), and at T4 median between-class variability had increased to 40% (IQR 22% to 49%). For example, in the primary school with the lowest seroprevalence at T4 (Figure 1i, leftmost bar), had classes with a minimum of 0% and a maximum of 40% seropositive subjects, thus a between class variability of 40%. Results were similar using a range of different inclusion criteria of classes in the analysis (e.g., 2 or 3 children tested per class; 2 participants per class and at least 2 classes per school; at least 3 participants and at least 50% participation in class). Furthermore, even within a single geographic district, seroprevalence varied widely between and within schools (Figure 2).

**Figure 1:**
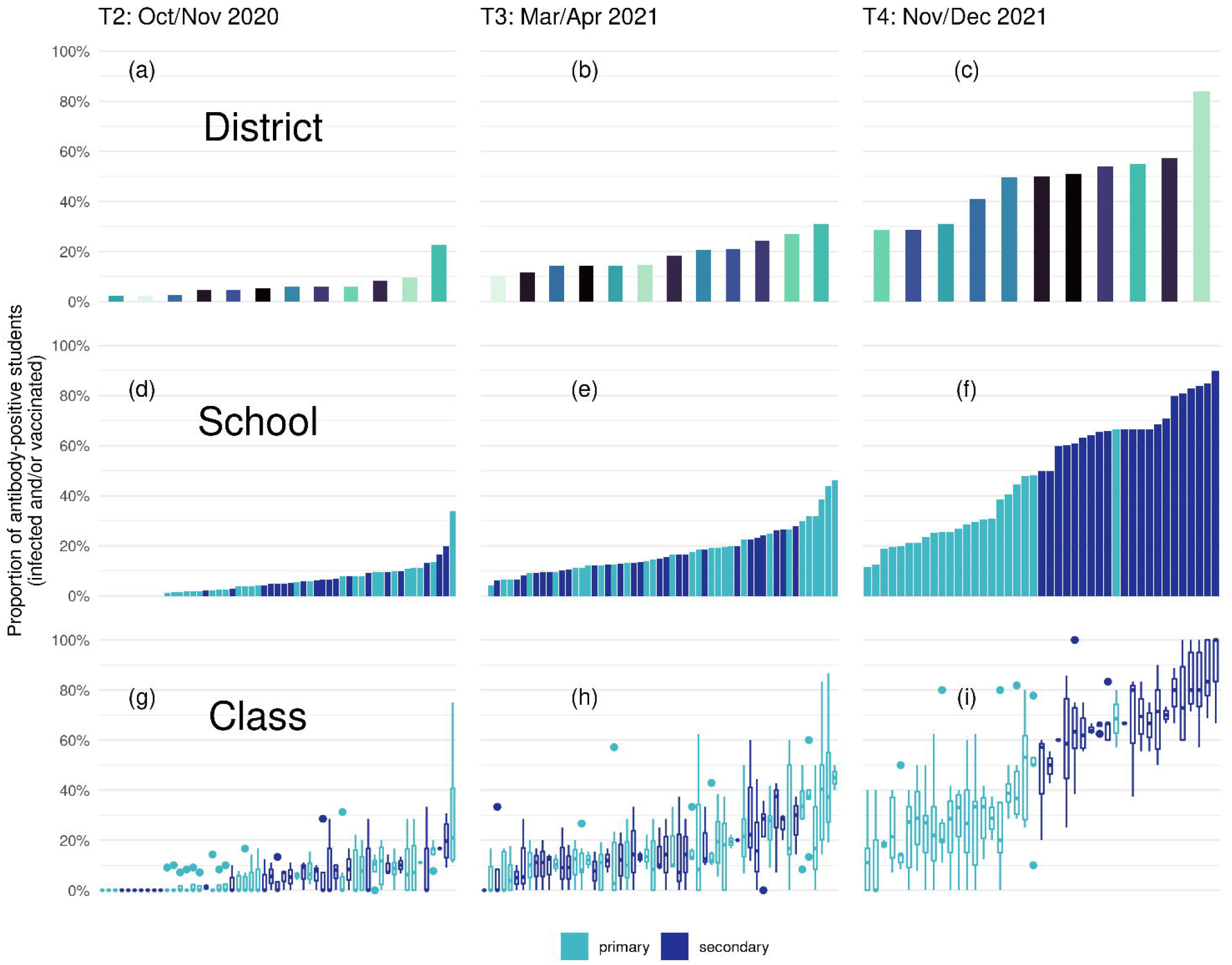
Proportion of ever-seropositive children in the canton of Zurich among districts (upper), schools (middle) and classes (lower) at T2 (October-November 2020), T3 (March-April 2021) and T4 (November-December 2021). Each district has an individual color in the upper panels, primary school children (grades 2-6) in light blue and secondary school children in dark blue (medium and lower panels). Boxplots in the lower panels denote median and describe variability of seroprevalence on a school level expressed as maximum seroprevalence in a class minus minimum seroprevalence and summarized as median (IQR). Whiskers of boxplots denote ± 1.5 standard deviation (SD).

**Figure 2.**
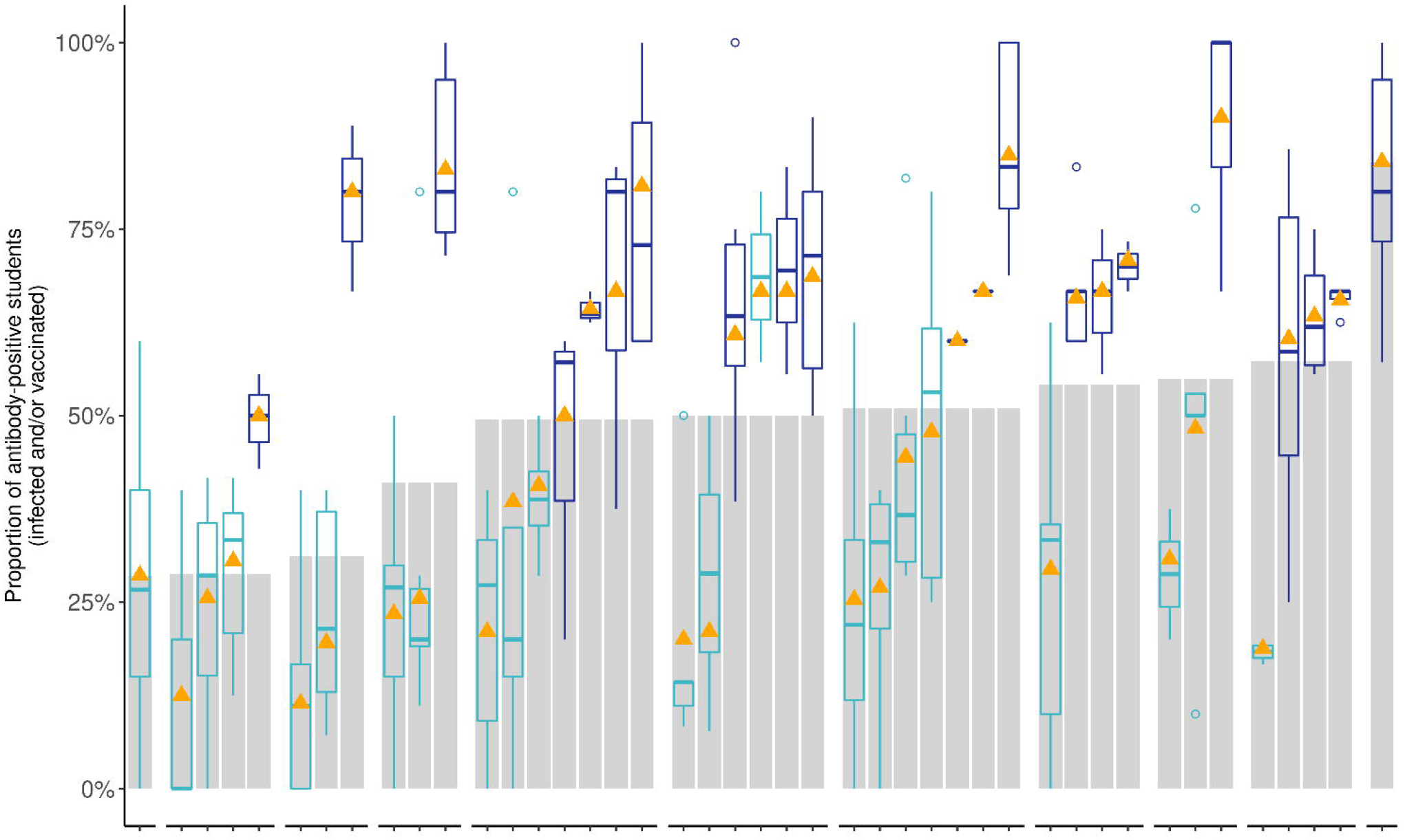
Variability of seroprevalence of children and adolescents at T4 (November-December 2021) among 11 districts (grey bars, mean), 43 schools (triangles, mean) and 200 classes (box-plots) within the same schools and districts. Along the horizontal axis, schools are displayed without names for reasons of confidentiality. Only classes where more than 5 children participated were included. Boxplots describe variability of seroprevalence on a school level, with middle bar indicating median seroprevalence, box representing IQR (middle 50% of classes) and whiskers the more extreme classes.

The overall explained variance was at most 25% (Figure 3, Table S1). Considering only unvaccinated children, explained total variance and variance explained by community, school and class were fairly similar but waned over time from T2 to T4. Also including the vaccinated children, explained variability increased markedly from T3 to T4 with most variability explained by school level, e.g., primary versus secondary school level.

**Figure 3.**
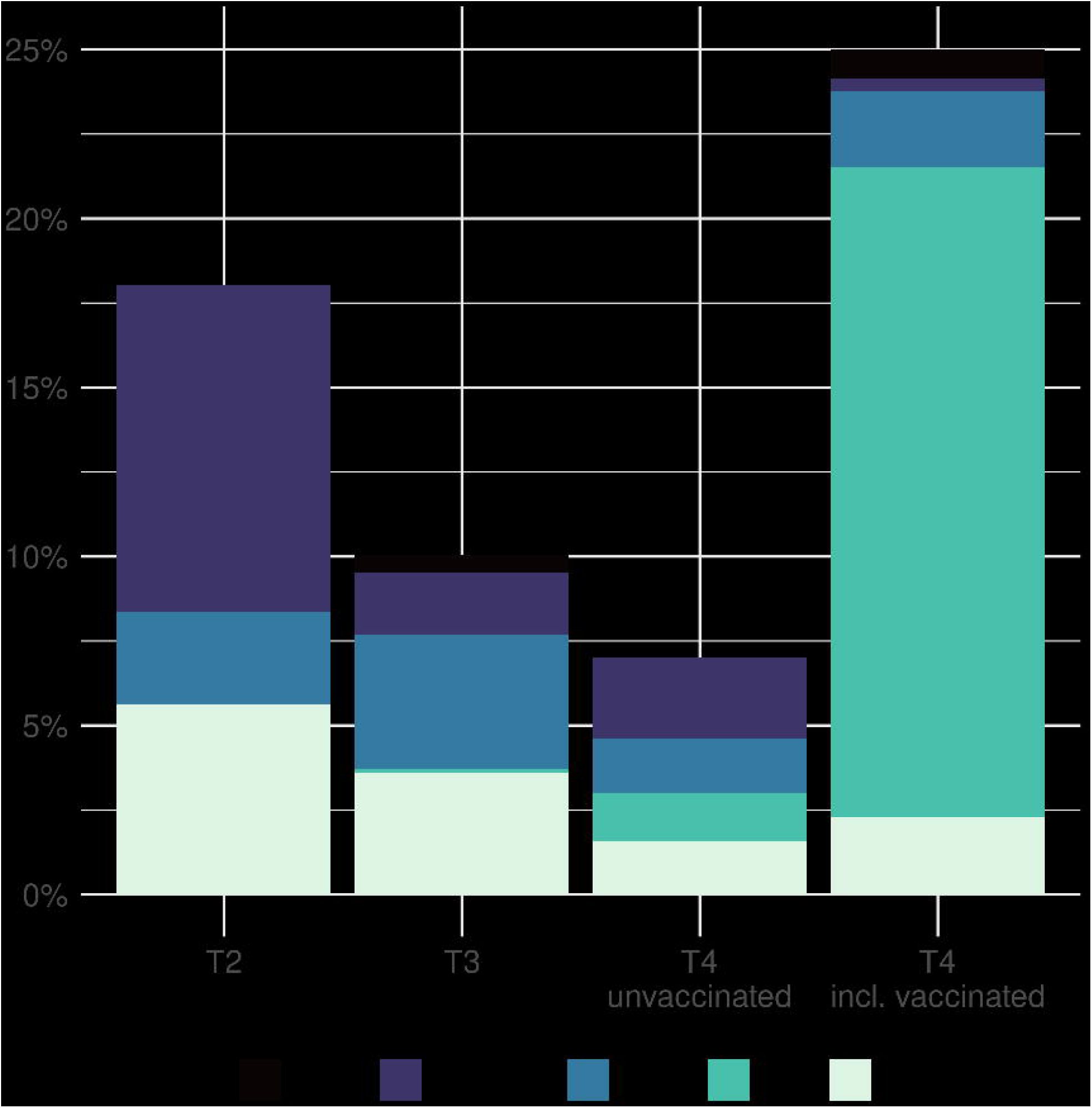
Composite variability in seroprevalence of children and adolescents at T2 (October-November 2020), T3 (March-April 2021) and T4 (November-December 2021) using variance partition coefficients. The total length of the bars shows the total proportion of variance in seroprevalence explained by class, school level (primary versus secondary), school (ID), community, and district. The remaining residual variation is attributable to unmeasured factors.

## Discussion

In this school-based cohort study of more than 1800 school children from randomly selected schools of the most populous canton in Switzerland, SARS-CoV-2 seroprevalence in children and adolescents increased from 6% in October/November 2020 to 46% in November/December 2021. The ratio of diagnosed to seropositive children in late 2021 (T4) of 1: 3.5 was similar to March – April 2021 (T3). s

These key points will be discussed in further detail below: Secondary school children had much higher seroprevalence rates than primary school children by the end of 2021, a difference which was not previously observed and only partially explained by higher vaccination rates. Variability of seroprevalence between districts, schools and classes within the same school increased substantially between the end of 2020 and the end of 2021 two- to fourfold. Explained variability over time decreased among unvaccinated subjects. This evolution mirrors an increasingly complex system of SARS-CoV-2 spread and transmission that is highly variable both in and outside of the school system, even in a relatively small part of the same country.

The increase in seroprevalence in children and adolescents in our study from 6% in October/November 2020 (T3) to 46% in November/December 2021 (T4) is consistent with other seroprevalence studies of Europe and the USA.^20-22^ Yet, between-country variability is substantial and is driven by multiple processes: vaccination status of children, adolescents and adults,^1^ some youth being vaccinated despite infection, heterogeneity of immune responses to SARS-CoV-2 with occasional failure to seroconvert after infection,^30^ and background community transmission.^31^

Until March-April 2021, primary school children consistently showed similar or slightly higher seroprevalence (<5%) than secondary school students, while this pattern clearly changed by November-December 2021. The 44% higher seropositivity rates among secondary school compared to primary school children are certainly explained by the introduction of the SARS-CoV-2 vaccination. In our analysis, 49% of those eligible to be vaccinated had completed the 2-dose regimen, and close to 100% of those were seropositive. It is not possible to determine which of these children also had antibodies due to infection from COVID-19. Considering that some of the vaccinated children were seropositive prior to vaccination (reflecting the seroprevalence of unvaccinated-infected children), the true seroprevalence in infected children would be expected to be higher than what was measured (extrapolating from the infected only children by about 10% to 15%). Not even N-antibodies (reflecting natural infection rather than vaccination) specifically found in infected individuals would have helped to estimate seroprevalence in infected children, as they wane after a few months, thus, much faster than our 8-month time window between T3 and T4.^32 33^ Even after excluding those vaccinated, the higher seroprevalence in secondary school children was also documented in other countries^1 12 33^ and was among the highest among age groups under the delta variant. Adolescents of this age group show a different social behavior than their younger counterparts, with more social contacts outside the home and schools setting, and they may also have been weary of the mitigation measures that were constantly in place since the beginning of the pandemic. In previous rounds, seroprevalence was higher in primary than secondary school level, but this was partially explained by differences in mask wearing policies.^16^ Masks were initially required only for the older age group which was associated with a 5% lower seroprevalence in adolescents and younger children in early 2021.^16^ The reversed trend in seroprevalence between primary and secondary school in late 2021 with the mask mandate changed to a general masking rule at school during most of the period between testing in 2021 reinforces this previous result.^16^ and could also be supported by a different social behavior of adolescents and a higher susceptibility compared to the younger age group in and outside school.^34^

Interestingly, explained variability was rather low and decreased over time when vaccinated children were excluded, but increased when all children, also the vaccinated children were considered. In the unvaccinated group, the dominant part of explained variance at T2 was the community where the child lived and his or her class.^17 24^ School became more important at T3,^16^ while at T4 neither community, nor school, school level or class explained much variability. With the vaccination of 48% of participating secondary school children, school level became the dominant factor, as the age of children in secondary school corresponded well to the age group where vaccination was available, 12 years and older. Even though the spread in school increased with more infectious variants, variation was not explained by school or class identity. Other factors such as the household or close contacts outside class and school could still be more important in the infection spread.^34^

From a public health perspective, Ciao Corona is unique as it repeatedly measures seroprevalence, an important measure to document the spread of infection in children and adolescents in the school setting, as well to assess the impact of children’s seroprevalence and vaccination on SARS-CoV-2 spread in schools. Ciao Corona is one of the few large studies reporting variation in seroprevalence over time in children within districts, schools, and school classes from randomly selected schools in a country where the general lock-down on a population level was mild and short (6 weeks in 2020), and school closure lasted only for 2 months at the start of the pandemic. We were able to perform 4 assessments covering the major SARS-CoV-2 variants (wild type at T1/T2, alpha in T3, delta in T4). The overall retention rate remained comparatively high through March/April 2021 (89 and 87%) although it decreased by T4 (34%), which mainly reflected decisions not to participate in T4 on a school rather than individual level. Use of serological testing implies that children with asymptomatic infections were also detected.

This study also has limitations. Due to the nature of serological testing, exact timing of infections cannot be determined. Therefore, examination of associated infections, in the sense of outbreaks or temporal clusters of infections, is not possible. We used a highly accurate serological test and adjusted for inaccuracy using Bayesian models on a population level, but it is not possible to avoid some false positive or false negative results on an individual level. Additionally, there were likely vaccinated children and adolescents who were also infected, and therefore some underestimation of seroprevalence of the infected only cohort is likely. Including these children also as infected would increase the seroprevalence estimates due to infection rather than vaccination. Moreover, the addition of Anti-N IgG antibodies that document natural infections SARS-CoV-2 also in vaccinated children may have helped to tease out the percentage of vaccinated children who also had a natural infection, although children seem to develop lower levels of anti-Nucleocapsid IgG antibodies with a faster decline than adults and the temporal sequence of natural infection and vaccination would still be missing.^30 35^ Participation bias in studies like Ciao Corona can occur at the individual level of the child, or on the class or school level; it can be balanced over time or differential non-participation at some time periods might have occurred. Due to the nature of repetitive testing with fear of venous blood sampling in mind, possible participation bias is unavoidable. Yet, we managed to have much higher participation rates than other similar studies (33 vs. 9%).^36^ Overall, we had comparable study populations of higher socio-economic state than the general Swiss population at each round, potentially leading to some underestimation of seroprevalence in each testing round as more disadvantaged populations show higher SARS-CoV-2 seroprevalence.^36 37^ Selective non-participation of children with known previous infections compared to previously seronegative children did not occur (e.g. 66 vs 62% of seropositive and seronegative children at T3 participated again at T4), but some misclassification could have taken place as antibody levels may wane over time.^38^ Yet, based on the relatively higher participation of secondary compared to primary school children and lower participation rates on the class level at T4 compared to previous rounds (see Table 2), some overestimation of seroprevalence is not excluded. On the school level, we examined the social index of the schools, a composite measure which reflects socioeconomic status (e.g., unemployment, immigrant population, parental support by the social system) of the location of the school provided by the Educational Directorate of Canton Zurich. The social index was representative for the canton of Zurich, did not change over time. and was comparable among those schools which participated or did not participate at T4. Because of a lower participation on the class level with about half of classes newly entering the study at T4 (with unknown previous serological status of these children) and a high number of vaccinated children in which concurrent natural infections could not be defined with our design, clustering as indication of intra-class or intra-school transmission could no longer be determined.

## Conclusion

We observed a large increase in seroprevalence from T2 to T4, especially from T3 to T4 following introduction of the vaccine for children 12 years and older. Up to T3, primary school children had higher seroprevalence, however at T4, secondary school children were more likely to be seropositive. This shift was in part due to introduction of the COVID-19 vaccine, but possibly also due to different behavior with more social contacts of older versus younger children outside schools and households. Variability in seroprevalence among districts, schools and classes was high and increased over time, even between different schools of the same district and among classes in the same school. Since this variability was not explained by school or class, other not captured factors (e.g., family members and other close contacts outside of the school setting) could still be more important in the spread of infection.

### Data sharing statement

Data is still being collected for the longitudinal cohort study Ciao Corona. Upon study completion in 2023, de-identified and potentially aggregated participant data, together with required data dictionaries, will be available on reasonable request by email to the corresponding author. The purpose and methods of data analysis will be evaluated by the study team first to ensure that it complies with the ethics approval.

## Data Availability

All data produced in the present study are available upon reasonable request to the authors.

## Contributors

SK and MAP initiated the project and preliminary design. SK, MAP, AU, TR, SRH developed the design and methodology. SK, TR, AU, AR, SR recruited study participants, collected, and managed the data. SRH performed statistical analysis and wrote the first draft of the manuscript. All authors contributed to the design of the study and interpretation of its results and revised and approved the manuscript for intellectual content. SK, SR, AR and SRH had access to and verified all underlying data. The corresponding author SK attests that all listed authors meet authorship criteria and that no others meeting the criteria have been omitted.

## Financial Disclosure

This study is part of Corona Immunitas research network, coordinated by the Swiss School of Public Health (SSPH+), and funded by fundraising of SSPH+ that includes funds of the Swiss Federal Office of Public Health and private funders (ethical guidelines for funding stated by SSPH+ will be respected), by funds of the Cantons of Switzerland (Vaud, Zurich, and Basel) and by institutional funds of the Universities. Additional funding, specific to this study is available from the University of Zurich Foundation.

## Competing Interests

All authors have completed and submitted the International Committee of Medical Journal Editors form for disclosure of potential conflicts of interest. No potential conflicts of interest were disclosed.

## Notes

### Competing Interest Statement

The authors have declared no competing interest.

### Author Declarations

The Ethics Committee of the Canton of Zurich, Switzerland (2020-01336) gave ethical approval for this work.

### Summary of Updates

Minor changes to the text regarding interpretation of differences between seroprevalence by school level.

