## Supplementary material for "Heterogeneous evolution of SARS-CoV-2 seroprevalence in school-age children: Results from the school-based cohort study Ciao Corona in November-December 2021 in the canton of Zurich" (from another version of this preprint: version 3, 2022-09-20)

Sarah R Haile PhD, Alessia Raineri MSc, Sonja Rueegg BSc, Thomas Radtke PhD, Agne Ulyte MD PhD, Milo A Puhan MD PhD, Susi Kriemler MD

**Table S1:** *Proportion of variance in seroprevalence explained by geographic district, community, school, school level and class, expressed using variance partition coefficient (percent of total variation explained by within cluster variation).*

|  | **Oct-Nov 2020**  (T2) | **Mar-Apr 2021**  **(T3)** | **Nov-Dec 2021**  unvaccinated*  (T4) | **Nov-Dec 2021**  vaccinated or unvaccinated*  (T4) |
| --- | --- | --- | --- | --- |
| Regional |  |  |  |  |
| district | 0% | 0.5% | 0% | 0.9% |
| community | 9.7% | 1.9% | 2.4% | 0.4% |
| School |  |  |  |  |
| School ID | 2.7% | 4.0% | 1.6% | 2.3% |
| School level | 0% | 1.0% | 1.4% | 19.2% |
| class | 5.6% | 3.6% | 1.6% | 2.3% |
| Total | 18.0% | 10.0% | 7.0% | 25.1% |

*T4 unvaccinated denotes the cohort with only unvaccinated children, T4 vaccinated or unvaccinated denotes the full cohort with vaccinated and unvaccinated children. School ID denotes the individual school.

**Figures S1** Average daily cases per 100,000 inhabitants in the canton of Zurich (blue) and across Switzerland (red).

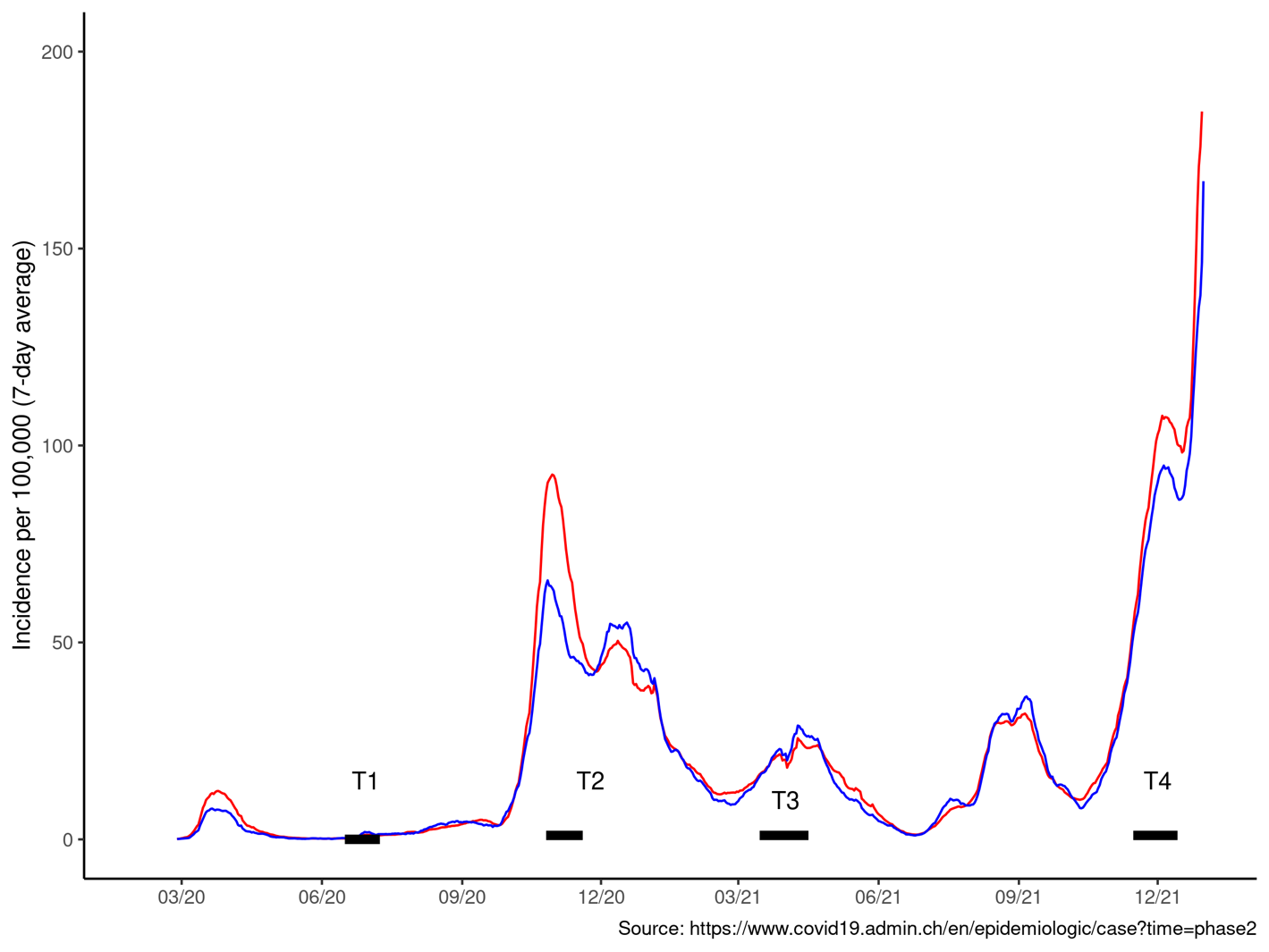
